# Novel coronavirus (COVID-19) Outbreak in Iraq: The First Wave and Future Scenario

**DOI:** 10.1101/2020.06.23.20138370

**Authors:** Adil R. Sarhan, Mohammed H. Flaih, Thaer A. Hussein, Khwam R. Hussein

## Abstract

The first patient with COVID-19 was reported in Iraq on 24 February 2020 for the Iranian student came from Iran. As of 24 May 2020, the confirmed cases of COVID-19 infections reached 4469, with 160 deaths and 2738 patients were recovered from the infection. Significant public health strategies have been implemented by the authorities to contain the outbreak nationwide. Nevertheless however, the number of cases is still rising dramatically. Here, we aim to describe a comprehensive and epidemiological study of all cases diagnosed in Iraq by 24 May 2020. Most of the cases were recorded in Baghdad followed by Basra and Najaf. About 45% of the patients were female (with 31% deaths of the total cases) and 55% were male (with 68% deaths of the total cases). Most cases are between the ages of (20-59) years old, and (30-39) years are the most affected range (19%) Approximately (8%) of cases are children under 10 years old. Iraq has shown a cure rate lower than those reported by Iran, Turkey and Jordan; and higher than Saudi Arabia and Kuwait. Healthcare workers represented about (5%) of the total confirmed cases. These findings enable us to understand COVID-19 epidemiology and prevalence in Iraq that can alert the our community to the risk of this novel coronavirus and serve as a baseline for future studies.

## Introduction

Coronavirus is one of the major viruses which primarily affecting the respiratory system in human (1). However, Coronaviruses have been also diagnosed in animals and can cause a range of severe diseases such as gastroenteritis and pneumonia (2,3). Previous coronavirus outbreaks have been reported, including severe acute respiratory syndrome (SARS-CoV) and Middle East respiratory syndrome (MERS-CoV), which is described as a significant public health threat (4). In 2002, coronavirus infections (SARS-CoVs) spread in Guangdong, south China, causing high fever, breathlessness and pneumonia, and rapidly spread to various regions around the world. The infection has spread in 26 countries, resulting about 8096 cases and 774 deaths (5,6). Whereas MERS-CoV was first detected in Saudi Arabia in 2012. The disease has mild respiratory symptoms that can lead to acute respiratory syndrome and death. 2494 cases were infected by the virus, of which 858 died in more than 25 countries (7–9). On December 2019, Atypical unkown pneumonia was first recorded in Wuhan city, Hubei province. Patients have showed high fever (more than 38 C°, dry cough, malaise, and breath difficulties. The infection has been linked to the seafood market of Wuhan, China and named COVID-19 (10–12). It spread rapidly to other Far East Asian nations, then to the Middle East and Europe. In severe cases the disease causes pneumonia, septic shock, metabolic acidosis and bleeding (13). The incubation period has been estimated from 5 - 14 days and may vary from patient to patient according to age and infection history (14).

Several studies have revealed that COVID-19 can be transmitted between humans via nasal droplets and direct contact in both symptomatic and asymptomatic patients (15–17). No vaccine or effective medication currently available to prevent or cure COVID-19 infections; however, some preventive health measures can help to resolve primary complications in patients (18). On 16 March 2020, the disease affected more than 150 countries and territories around the world. Over the past few months there has been a significant increase in COVID-19 cases. In Iraq, the first confirmed case of COVID-19 has been reported in Najaf province for the Iranian student came from Iran on 24 February 2020, followed by 4 cases from one family in Kirkuk province on 25 February, they have also a travel history to Iran. An additional case was recorded on 27 February in Baghdad, for a patient who recently visited Iran. (19). 74 confirmed cases and 8 fatalities have been reported across Iraq as of 12 March 2020 (20). The confirmed cases jumped to 1415 on 16 April 2020, with 78 fatalities were recorded (21). By 24 May 2020, the confirmed cases of COVID-19 reached 4469 and reported 160 deaths, while 2738 patients recovered from the infection (22). Here, we aim to describe a comprehensive, epidemiological study of all cases diagnosed in Iraq by 24 May 2020. We hope our study will alert the community to the risk of this novel coronavirus, in order to prevent a second wave of the virus infections.

## Methods

This study was an exploratory and descriptive analysis of all COVID-19 confirmed cases, diagnosed in Iraq as of 24 May 2020. We obtained epidemiological and demographic data for all COVID-19 cases that were reported from 24 February 2020 to 24 May 2020 by the Iraqi Ministry of Health, Directorate of Public Health. Sex-disaggregated data about COVID-19 was obtained from Global Health 50/50. We collected data for global COVID-19 cases from ProMED, WHO, and CDC reports.

Case fatality rates (CFR) were calculated as the total number of deaths (td) divided by the total number of cases (tc), represented as a percentage (23,24).

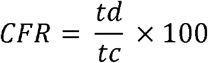

While the incidence rates (IR) were calculated as the total number of COVID-19 confirmed cases (tc) divided by the population (p) of each province times 100,000 (25).

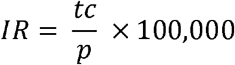

This study was evaluated according to the reporting guidelines for Strengthening the Reporting of Observational Studies in Epidemiology (STROBE) (26).

## Results

### Demographics and the distribution of COVID-19

Overall, 4469 cases were confirmed with COVID□19 infection and showed a substantial cumulative rise since the first case was confirmed on 24 Feb 2020 until 24 May 2020. (Figure 1A and B). In comparison, Anbar, Ninewa, Diwaniya and Sala-Aldin have reported the lowest number of COVID□19 confirmed cases. The most cases were registered in Baghdad (2233 cases) followed by Basra (747 cases) and Najaf (318 cases) (Figure 1A). In contrast to other cities, the fatalities of COVID-19 infection in Baghdad was high (97 deaths), followed by Basra (18 deaths) and Kerbala with 8 deaths (Figure 1C). On the other hand, Erbil (Kurdistan region, north Iraq) registered only one death for 242 confirmed cases. In addition, Figure 1D shows the breakdown of confirmed cases and deaths by gender across all cities. Approximately 45% of patients were female and 55% were male. While the death rates were 68% for males and 31% for females.

**Figure 1:**
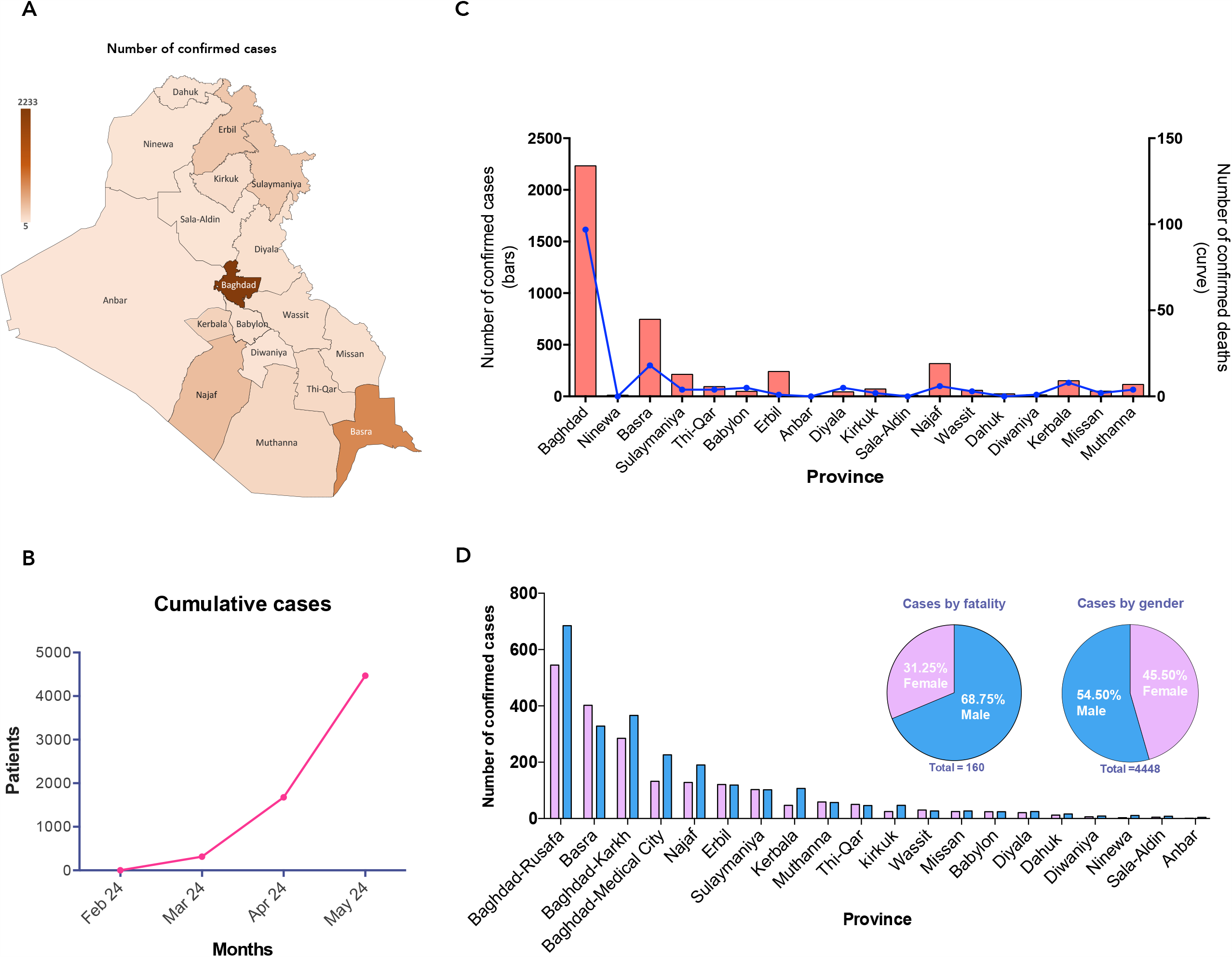
Demographics and the distribution of COVID-19. **A**. The number of all confirmed cases reported for all provinces in Iraq and represented as color-coded heatmap. **B**. Cumulative cases that have been reported between 24 Feb 2020 and 24 May 2020. **C**. Direct comparison between the total number of all cases with the total number of deaths in all provinces. **D**. Cases and deaths by gender across all cities.

### COVID-19 cases and indicators by province and age

Data revealed that most cases are between the ages of 20-59, with those aged 30-39 years being the most affected range (19 %). Such age groups are the ones that are most likely to have a high chance of participation in different sectors of work (Figure 2A). Data have also shown that around 8% of overall confirmed cases are children under 10 years old (Figure 2A). As can be seen from the Figure 2A, there is a clear trend of increasing in the fatality rate with ages between 60-80 years old reaching the max in patients with above 80 years old (24%).

**Figure 2:**
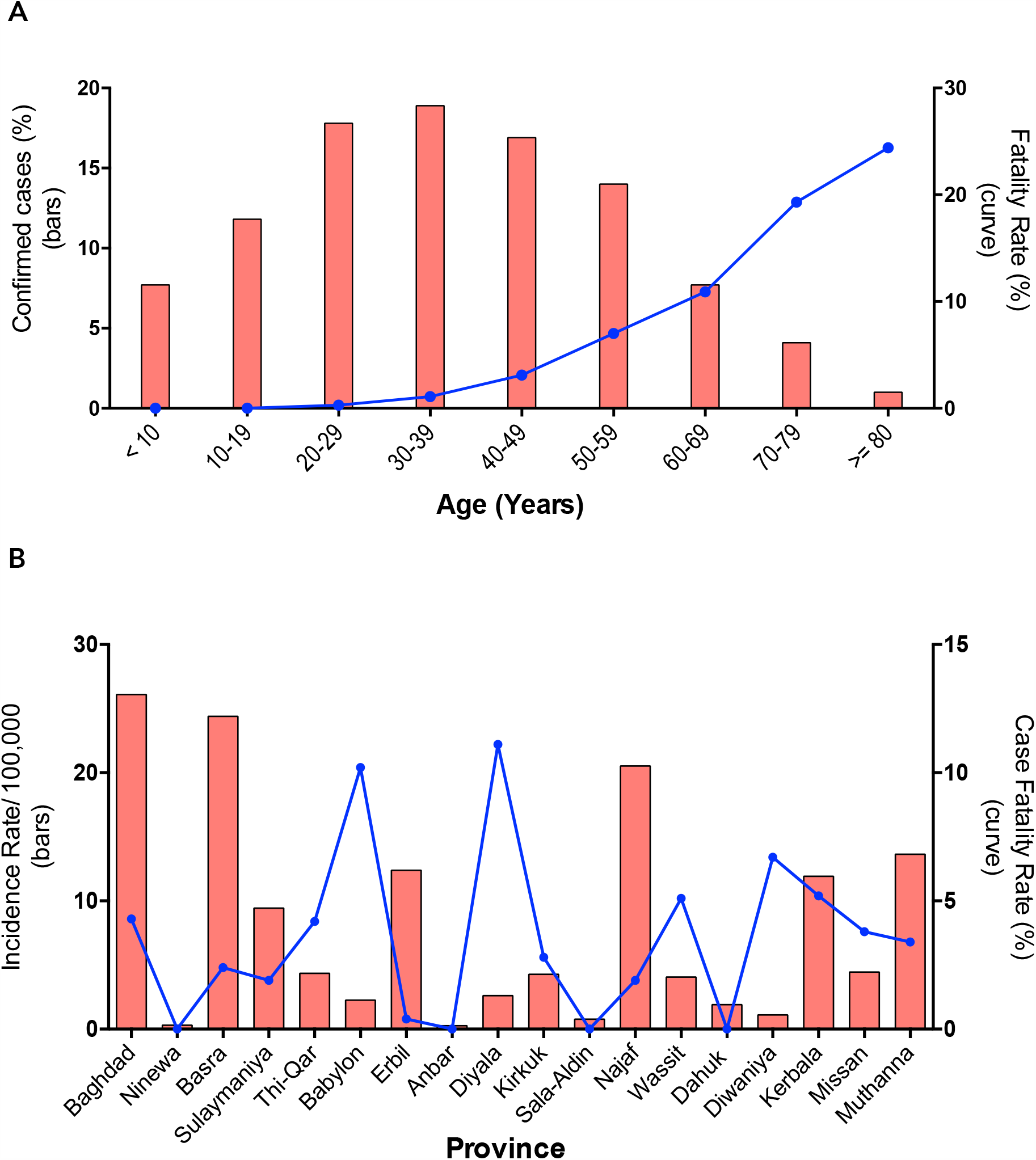
COVID-19 cases and indicators by province and age. **A**. The number of cases and the fatality rates across ages. **B**. Represents the incidence and case fatality rates in all provinces.

The provinces are widely varied in their incidence and case fatality rates (CFR) (Figure 2B). The incidence rate, which is the number of new cases in the population over a given period, has been calculated in all provinces. A higher incidence rate can be seen in Baghdad (26.09) followed by Basra with (24.39) and Najaf with (20.52) (Figure 2B). The high incidence rate in those large cities could be explained by the high number of confirmed cases. Although Anbar, Ninewa and Sala-Aldin demonstrated the lowest incidence rates ranging between (0.27-0.77). On the other hand, despite these findings on incidence rates, the case fatality rates were higher in Diyala (11%) and Babylon (10%), followed by Diwaniya (7%) (Figure 2B).

Among the confirmed cases of COVID□19, cure rates have shown a promising trend in some cities (Figure 3). For example, Najaf showed (94%) cure rate of the 318 confirmed cases. Erbil and Missan showed a cure rate of (87%). Despite this, Baghdad represented the lowest cure rate with (42%).

**Figure 3:**
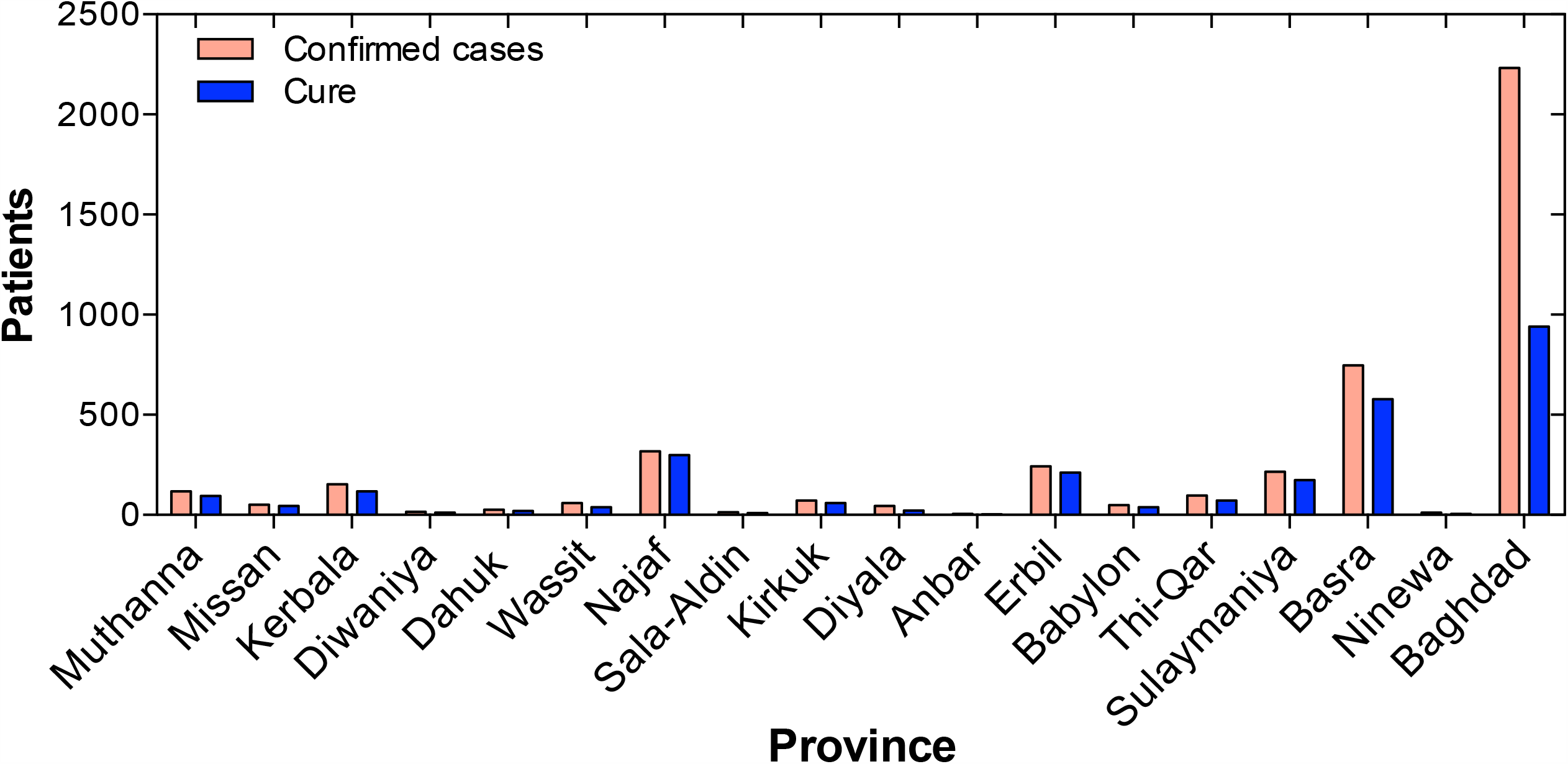
Confirmed COVID□19 cases and cure. Comparison among the number of cases in all provinces with the number of cure.

### Healthcare workers

By 24 May 2020, a total of 207 healthcare workers had been infected with COVID-19 (Figure 4). Represented about (5%) of the total confirmed cases in Iraq. The most interesting aspect of this data is that around (80%) of these infections were located in Baghdad, Najaf, Basra and Sulaymaniya. The majority of the infected health workers were nursing staff (60%), followed by physicians (30%) (Figure 4). These two groups have a greater chance of being in contact with infected patients.

**Figure 4:**
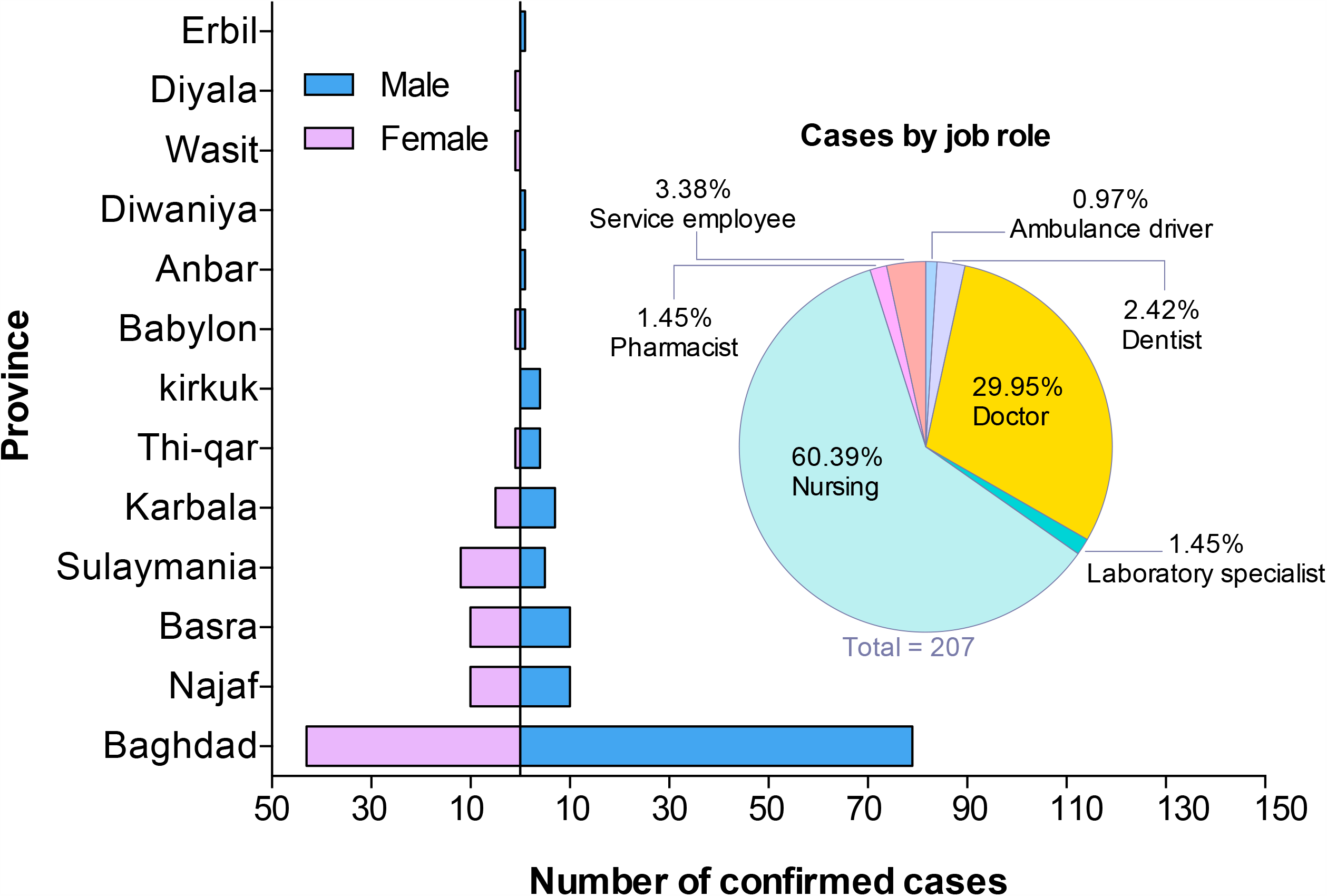
Healthcare workers who were infected with COVID-19. COVID-19 infections among healthcare workers and were classified according to gender and job role.

### COVID-19 cure and fatality rate in Iraq and neighboring countries

Iraq has shown a (61%) cure rate lower than those reported by Iran, Turkey and Jordan (Figure 5A). This figure is, however, higher than the reported cure rate for other neighboring countries. On the other hand, Iraq’s fatality rate was (4%) lower than Iran, which is indicated the high rate of fatality (5%). Saudi Arabia, by contrast, showed the lowest fatality rate (0.5%) (Figure 5B).

**Figure 5:**
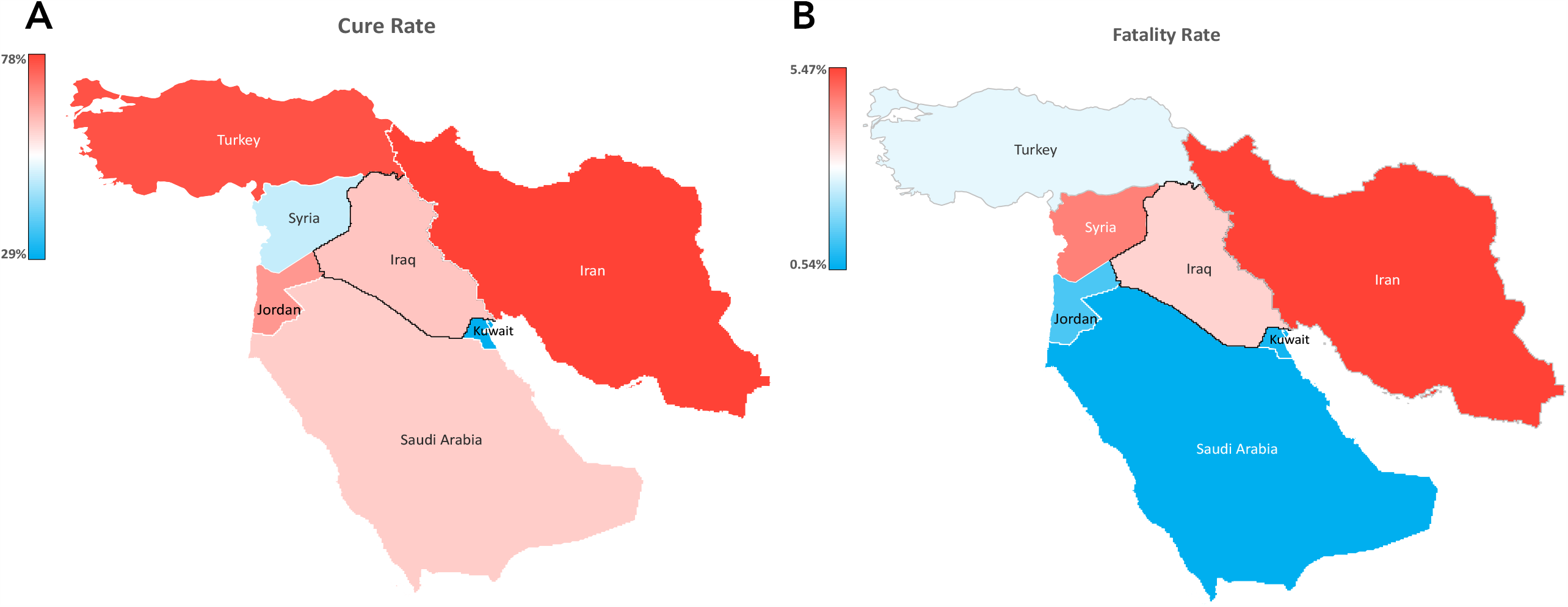
COVID-19 cure and fatality rate in Iraq and neighboring countries. **A**. The cure rate in Iraq and neighboring countries. **B**. The fatality rate in Iraq and neighboring countries.

## Discussion

The recent COVID-19 is a continuing pandemic that creates confusion and panic around the world. According to the genomic studies, the virus is considered to originate from infected bats, and can be transmitted to humans and animals, as well as to humans (27). It was found to have a positive-sensed, single-strand beta-coronavirus RNA virus similar to Bat Coronavirus (BatCoV RaTG13) with 88% identity (27,28). By 24 May 2020, an astounding 5,230,755 confirmed COVID-19 cases were reported worldwide, with 339,588 fatalities.

In Iraq, as of 24 May, 4469 COVID□19 confirmed cases, 160 deaths and 2738 patients who have recovered from the virus and discharged from hospitals. In the last few weeks, the number of confirmed cases has risen by more than 70%, from 1677 cases on 24 April to 4469 on 24 May 2020. Unfortunately, while we were writing this manuscript, the number of confirmed cases increased dramatically from 4469 on 24 May to 15,414 on 10 June with 426 deaths. One possible explanation for this increase could be that during this period (24 May-10 June) people celebrated Eid Al-Fitr after fasting in the holly Ramadan, which exhibited many gatherings for all family members and friends. Another possible explanation is that authorities has generally relaxed enforcement of the stringent curfews and movement restrictions which have been in place for the past several weeks. Partial lockdowns are currently in force in order to limit the spreading of the virus. It’s worth notice that the total cumulative number of COVID-19 cases increasing significantly, so this is could be a sign that a country will faces a second wave of virus infections. Furthermore, the number of confirmed cases varies across provinces. The highest number of cases were recorded in Baghdad and Basra compared to other cities due to the fact that these provinces have a large population with many factories and shopping malls that could not follow the instructions of the health authorities to contain the spread of the virus. The current study also found 54% of confirmed cases were males, and 45% were females (male: female ratio = 1.2:1). In addition, the fatality rates were higher in male (68%) than female (31%). These data seem to be consistent with other studies which found that men with COVID-19 infection are more likely to have serious consequences and death (29,30). Differences between males and females in getting infection with COVID-19 remain unclear. Countries like Italy, Scotland, Switzerland, Sweden and Belgium have reported a higher percentage of cases among women according to Global Health 50/50 (31). While the rates of infection among men appear to be much higher in Iran, Costa Rica, Thailand, Greece, Pakistan and Mexico.

In our study, data showed all ages are susceptible to infected with COVID-19 and the highest range was between 30-39 years old. Consistent with other studies (32–34), this research found that fatality rates were higher in patients with ≥60 years old. COVID-19 negative outcomes tend to be related to comorbidity including diabetes, obesity, cardiovascular and lung diseases. These factors may account for high death rates in older patients. Despite this, the majority of provinces, apart from Baghdad, showed promising cure rates.

As our healthcare workers continue to fight COVID-19, they get infected with the virus. The authorities start to be concerned that this could make a significant contribution to increasing the number of cases among healthcare workers and could then transmit the virus to their families and loved ones. Overall, 207 Healthcare workers were tested positive for COVID-19 representing (5%) of the total confirmed cases in Iraq. What stands out in the data is that (78%) of these cases were in Baghdad, Najaf and Basra. Cure and fatality rates have shown variation when comparing Iraq with neighboring countries. Iran reported the highest rates of cure and fatality at the same time. This inconsistency may be due to the fact that each country has different management protocols, case reporting and the number of tests carried out on each day. Some countries may not report mild cases if they do not need treatment or if they have been treated at home. There are some limitations to this study. First, We were unable to evaluate the clinical characteristics of confirmed cases since these data were not accessible at the moment of analysis. Second, we did not have any information as to whether or not these patients, especially elderly patients, have comorbidities. A further study with more focus on the comorbidities and the severity of COVID-19 in elderly is therefore suggested. Taken together, these findings contribute to our understanding of the epidemiology and prevalence of COVID-19 in Iraq, in several ways, which may be useful and provide a basis for future studies.

## Data Availability

The data supporting the conclusions of this article will be made available by the authors.

